# Forecasting COVID-19 new cases in Algeria using Autoregressive fractionally integrated moving average Models (ARFIMA)

**DOI:** 10.1101/2020.05.03.20089615

**Authors:** Belkacem Balah, Messaoud Djeddou

## Abstract

In this research, an ARFIMA model is proposed to forecast new COVID-19 cases in Algeria two weeks ahead. In the present study, public health database from Algeria health ministry has been used to build an ARFIMA model and used to forecast COVID-19 new cases in Algeria until May 11, 2020.

**Background:** The aim of this study is first to find the best prediction method among the two techniques used and type of memory, either short or long, of the model constructed for the daily confirmed cases in Algeria, then make forecasts of the confirmed cases in the fifteen next days.

**Methods:** This study was conducted based on daily new cases of COVID-19 that were collected from the official website of Algerian Ministry of Health from March 1, 2020 to April 26, 2020. Auto Regressive Integrated Moving Average (ARFIMA) model was used to predict the trend of confirmed cases. The evaluation of the fractional differentiation parameter (*d*) is carried out using OxMetrics 6 software.

**Results:** The ARFIMA model (0, 0.431779, 0) build for Algeria, has a long memory and an upward trend over the next fifteen days and which coincides with the holy month of Ramadhan.

**Conclusions:** The forecasted results obtained by the proposed ARFIMA model can be used as a decision support tool to manage medical efforts and facilities against the COVID-19 pandemic crisis.

## Background

The statistical prediction models can be considered as tool in forecasting a pandemic situation. The COVID-19 new cases in Algeria time series will be analyzed and predicted via the use of known forecast models such as the ARFIMA (Autoregressive Fractionally: (AR), order of fractional differentiation: (d), Moving Average: (MA)) model. This model used to describe a process that evolves over time. The values of (AR) and (MA) are calculated using the graph of the autocorrelation function (ACF), and the graph of partial autocorrelation (PACF).

In this article, we build an ARFIMA model, and study the behavior and analyze the type of forecast memory (long or short) of this model using COVID-19 new cases in Algeria using two techniques:

1. Maximum likelihood.
2. Nonlinear least squares.

Long memory models have become increasingly popular as a tool for describing time series. Long-term memory (long-term correlation) in time series is mainly assessed using the exponent from Hurst [1–2–3].

The ARFIMA model was proposed by Granjer and Joyeux (1980) [4–5–6], and since then they draw particular attention to the class of ARFIMA models by their flexibility to model many real situations by estimating the parameter *d* [7].

## Methods

### Data sources

The daily COVID-19 new cases in Algeria were collected from the Ministry of Population Health and Hospital Reform official website from March 1, 2020 to April 26, 2020. A time series of 57 days is constructed with a total of 3382 cases, an average of 58 cases per day, and a standard deviation of 50 cases to develop the ARFIMA model.

### ARFIMA models

Hamid et al. [8] recommend that data used should follow a normal distribution, In order to verify this recommendation, the data set is tested using Jarque-Bera normality test, based on the distribution of a combined measure of asymmetry and kurtosis.

The stationary of the time series was verified using the Augmented Dickey and Fuller test.

Residuals analysis is the most important step to validate the constructed model. In order to check if the residuals are white noise, a Ljung-Box test is applied to the models obtained to verify the effect of homoschedastic, this test remains is a decisive tool for the adaptability of constructed model.

Hurst (1951) [3] proposed a useful statistical method for understanding the properties of a studied time series., it is often used to analyze long-term time series correlations [9–10]. Depending on the H value, it is possible to determine if the sequence is completely random or if there is a trend component. The trend component is expressed as persistent or anti-persistent, indicating the change in the number of daily infected cases recorded from the past compared to the future.

The value H ≠ 0 implies the existence of long-range correlations and corresponds to the so-called fractional Brownian motion.

H = 0, the autocorrelations are zero and the spectral density is constant and positive. There is therefore no long-term dependence on the process.

In particular, there is persistence for H > 0.5, and for H < 0.5 anti persistence (is called by Mandelbrot Joseph effect) it corresponds to alternations of rise and fall in the process [10–11–12].

If 0.5 < H <1, the autocorrelations are all positive and decrease hyperbolically towards zero, the spectral density exhibits a pole at zero frequency, the series presents all kinds of non-periodic cycles and the process has a persistent form of long memory whose explanation is different from that of short-term persistence.

If 0 < H <0.5, the autocorrelations alternate sign and the spectral density, zero at zero, is dominated by the high frequency components. The process is anti-persistent: rising phases tend to be followed by falling phases. According to [3]; the Hurst exponent is given by Eq. (1) below:

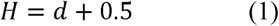

## Results

Jarque-Bera normality test (Fig.1) applied to the time seriese, shows that the series of COVID-19 new cases follows a normal distribution and the risk of rejecting the null hypothesis H_0_ is equal to 19.4%.

**Figure 1:**
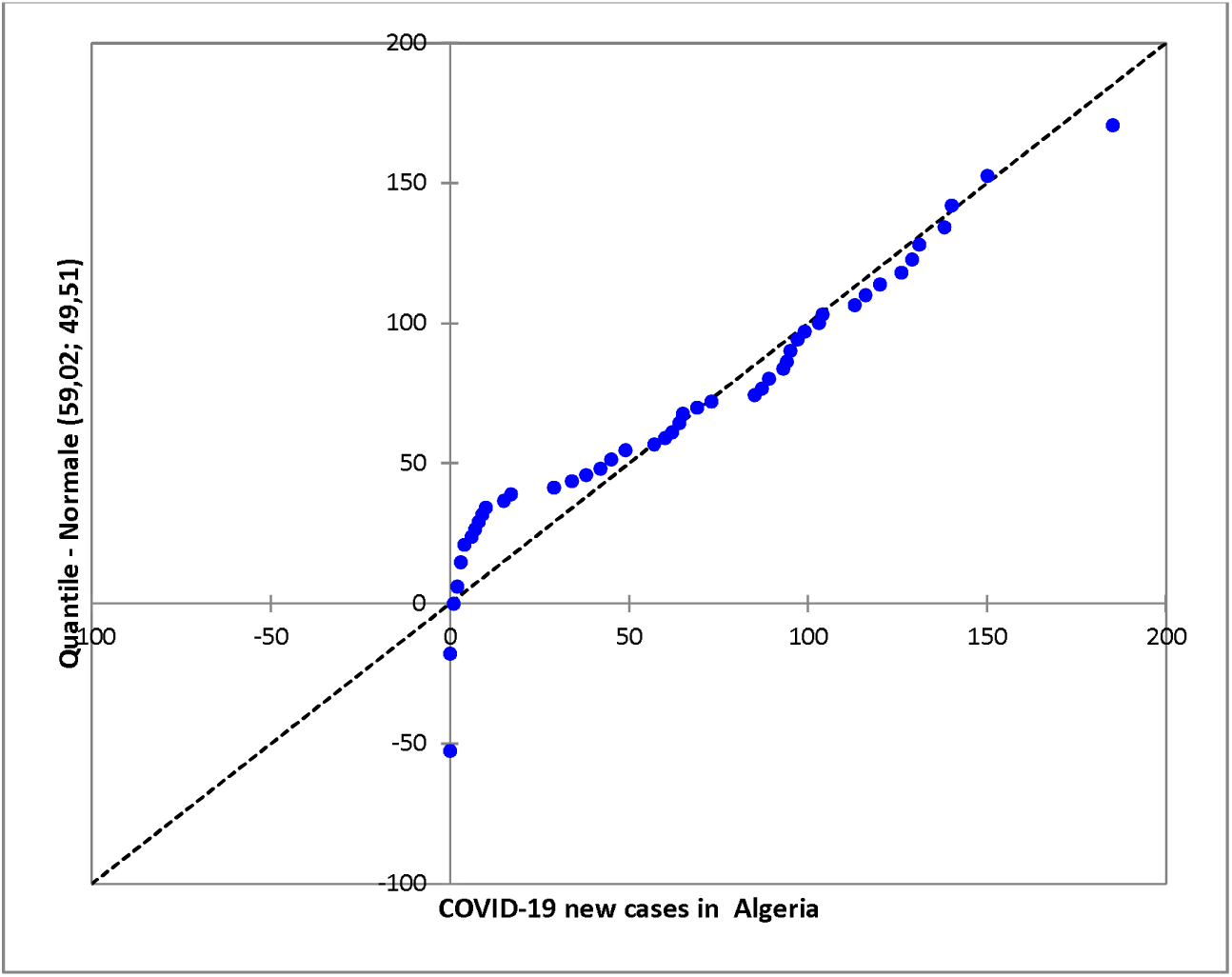
Quantile-quantile plot of COVID-19 new cases time series in Algeria.

PETTITT test was applied on the COVID-19 new cases time series, results showed that data series is not homogeneous, and has an increasing tendency in stairs. The first average is about 17 persons and the second average is 102 persons with March 30, 2020 as the break date (see Figure 2). Analysis of the data with Augmented Dickey-Fuller test (ADF) is proof of acceptance of the null hypothesis (H0) and the risk of rejection of the alternative hypothesis (Ha) is less than : 89% for the series of new cases of COVID-19. Therefore, the data series shows the absence of the unit root, therefore it is characterized as non-stationary.

**Figure 2.**
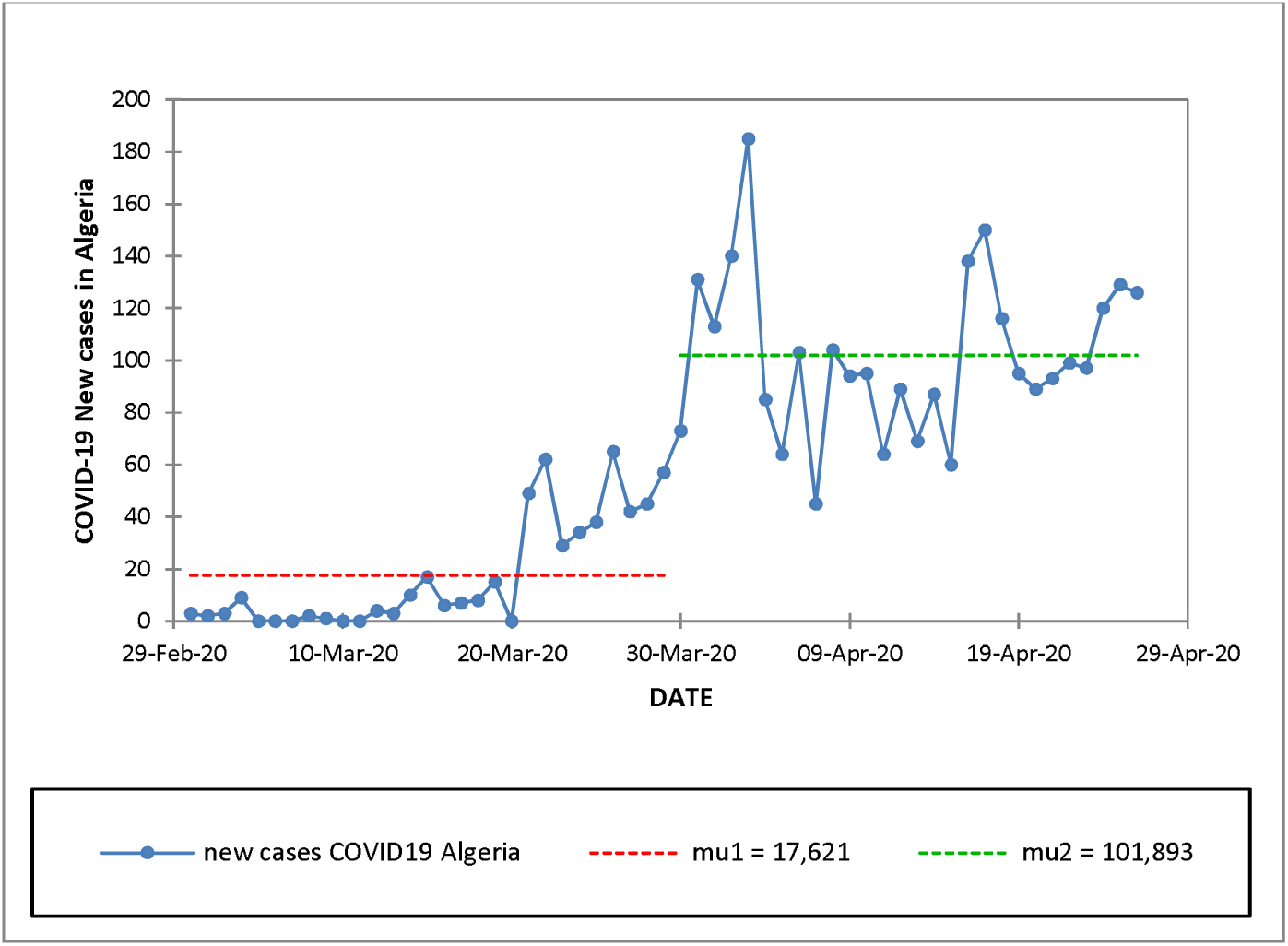
COVID-19 new cases in Algeria PETTIT test.

**Table 1.**
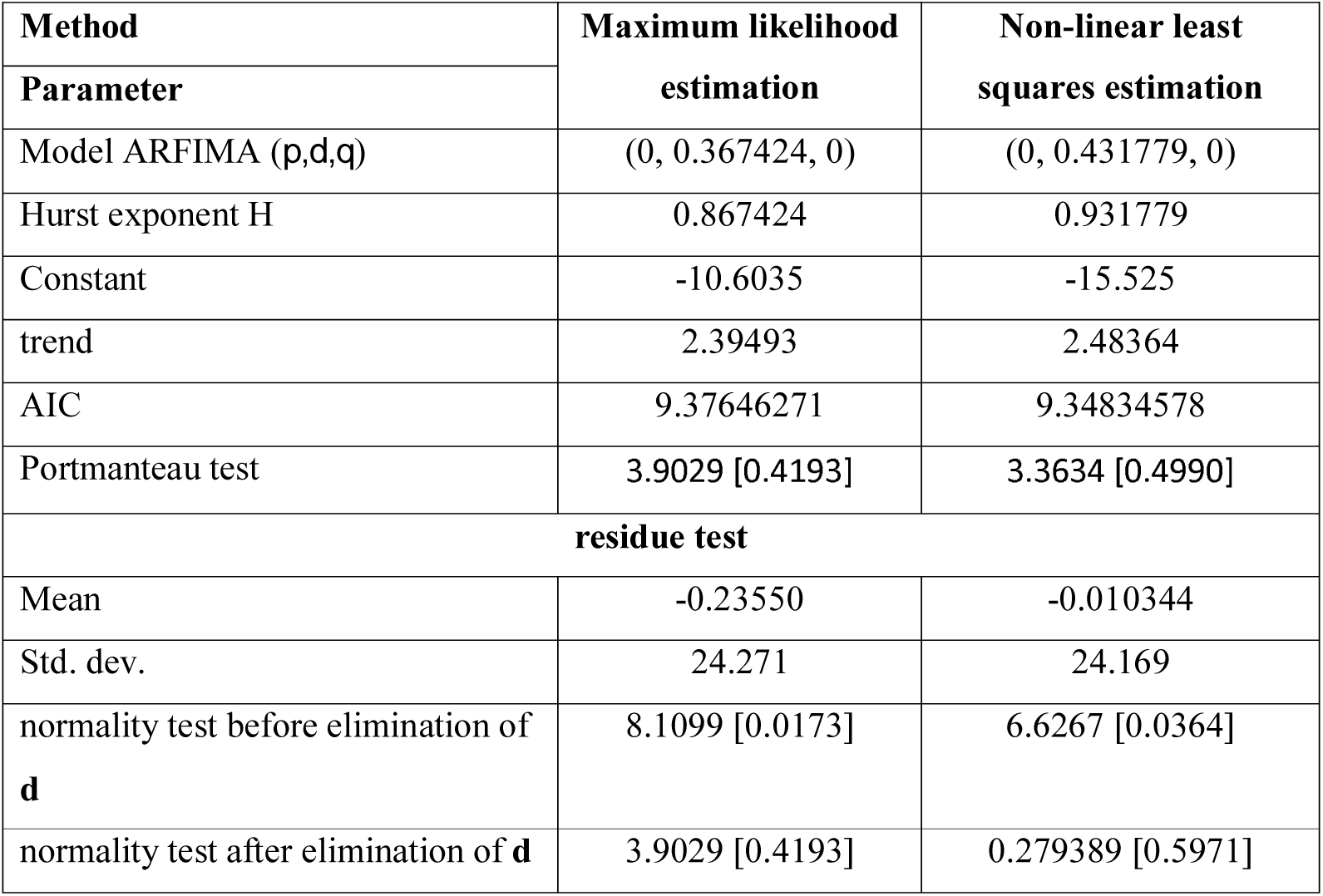
Modeling results of the COVID-19 new cases in Algeria using ARFIMA model with the corresponding Hurst exponent.

| Method | Maximum likelihood | Non-linear least |
| --- | --- | --- |
| Parameter | estimation | squares estimation |
| Model ARFIMA $(p,d,q)$ | (0, 0.367424, 0) | (0, 0.431779, 0) |
| Hurst exponent H | 0.867424 | 0.931779 |
| Constant | -10.6035 | -15.525 |
| trend | 2.39493 | 2.48364 |
| AIC | 9.37646271 | 9.34834578 |
| Portmanteau test | 3.9029 [0.4193] | 3.3634 [0.4990] |
| **residue test** |  |  |
| Mean | -0.23550 | -0.010344 |
| Std. dev. | 24.271 | 24.169 |
| normality test before elimination of **d** | 8.1099 [0.0173] | 6.6267 [0.0364] |
| normality test after elimination of **d** | 3.9029 [0.4193] | 0.279389 [0.5971] |

The selection of the best models follow three criteria’s:

1. Minimal residual standard deviation;
2. Akaike criterion (AIC);
3. Ljung Box (Portmanteau test) residual value ≥ the significance level of 5%.

Through these three criteria, the Non-linear least squares estimation method, whose ARFIMA model (0, 0.431779, 0) is better, has a long forecast memory until the next 15 days so that its Hurst coefficient equal to 0.931779 which is in the range of 0.5 to 1, and all of the results of this forecast are valid until May 11, 2020.

The Ljung-Box statistic in the table shows that the residuals of the ARFIMA model constructed is not a white noise process and the hypothesis of nullity H0 of homoscedasticity is accepted for a level of significance α = 49.9% of the series COVID-19 new cases. The sequential and partial autocorrelation diagrams are illustrated in Figure 3.

**Figure 3.**
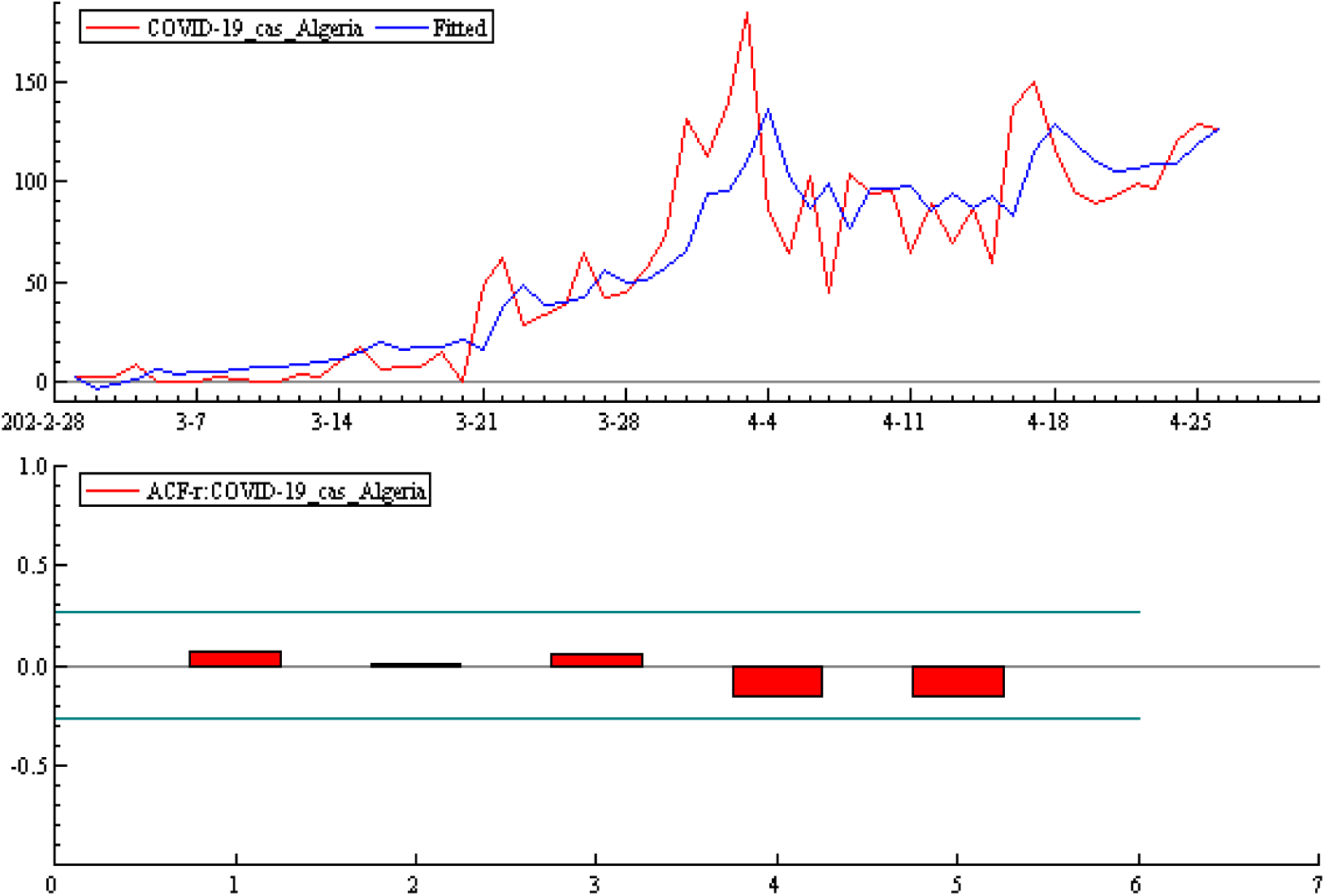
COVID-19 Confirmed new cases vs predicted COVID-19 new cases and partial autocorrelation using ARFIMA model

Figure 4 shows the forecast for the next 15 days (from April 26 to May 11) and we notice an upward trend over the period quoted above in this country, which is under pressure from the state to control the epidemic and the situation.

**Figure 4.**
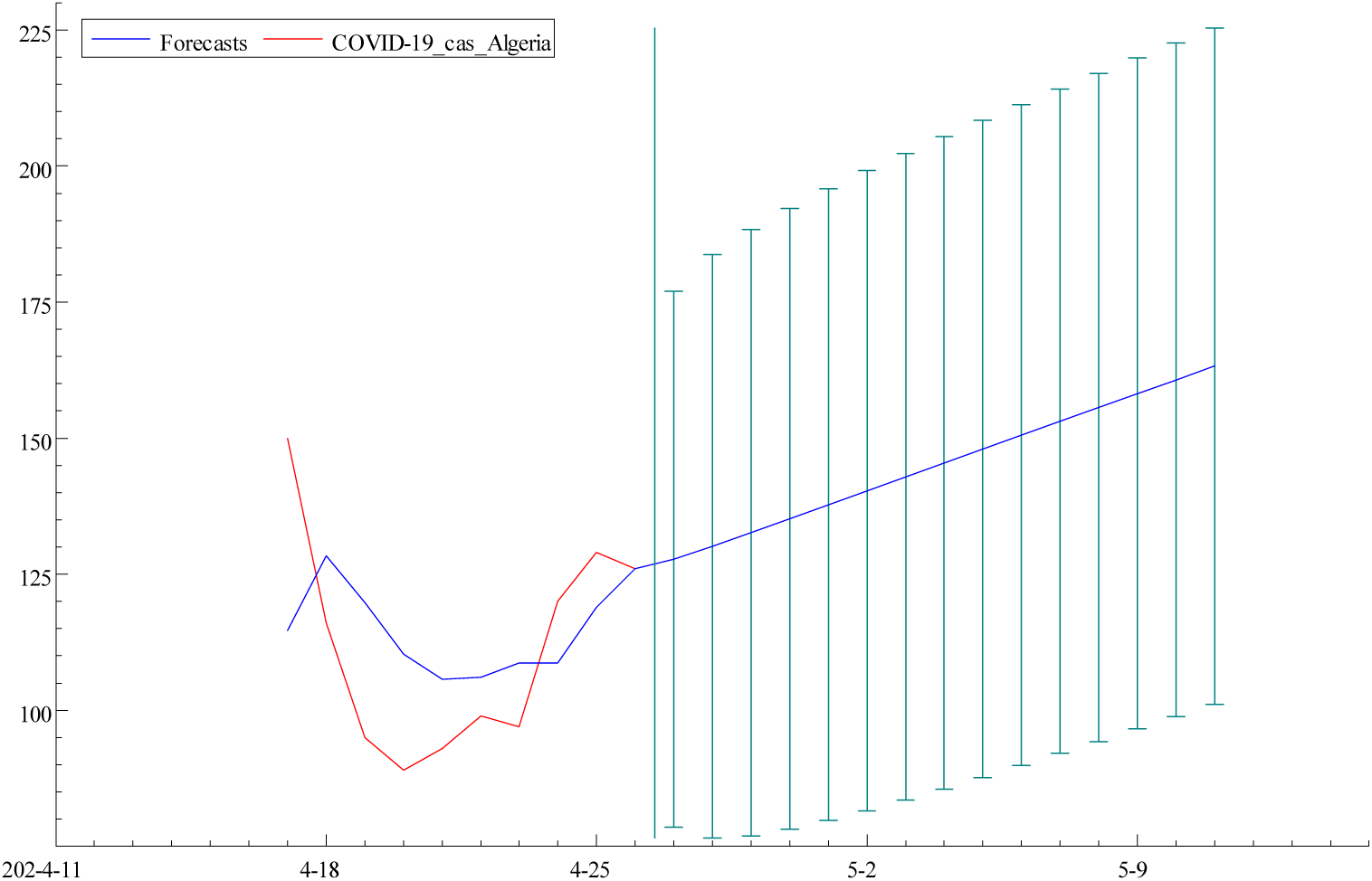
Forecasted COVID-19 new cases in Algeria until May 11, 2020 using ARFIMA model.

## Conclusion

Algeria’s efforts will have to focus on changing the increasing trend and could control the pandemic situation in near future. The quarantine should be in the front line program with the reduction of peoples travelling between departments.

The forecasted results obtained by the proposed ARFIMA model can be used as a decision support tool to manage medical efforts and facilities against the COVID-19 pandemic crisis.

## Data Availability

All the data were collected from the official website of Algerian Ministry of Health from March 1, 2020, to April 26, 2020. http://covid19.sante.gov.dz/carte/

https://fr.wikipedia.org/wiki/Pand%C3%A9mie_de_Covid-19_en_Alg%C3%A9rie

